# Integration of functional genomics and statistical fine-mapping systematically characterizes adult-onset and childhood-onset asthma genetic associations

**DOI:** 10.1101/2025.02.11.25322088

**Authors:** Xiaoyuan Zhong, Robert Mitchell, Christine Billstrand, Emma Thompson, Noboru J. Sakabe, Ivy Aneas, Isabella M. Salamone, Jing Gu, Anne I. Sperling, Nathan Schoettler, Marcelo A. Nóbrega, Xin He, Carole Ober

## Abstract

**Background:** Genome-wide association studies (GWAS) have identified hundreds of loci underlying adult-onset asthma (AOA) and childhood-onset asthma (COA). However, the causal variants, regulatory elements, and effector genes at these loci are largely unknown.

**Methods:** We performed heritability enrichment analysis to determine relevant cell types for AOA and COA, respectively. Next, we fine-mapped putative causal variants at AOA and COA loci. To improve the resolution of fine-mapping, we integrated ATAC-seq data in blood and lung cell types to annotate variants in candidate *cis*-regulatory elements (CREs).

We then computationally prioritized candidate CREs underlying asthma risk, experimentally assessed their enhancer activity by massively parallel reporter assay (MPRA) in bronchial epithelial cells (BECs) and further validated a subset by luciferase assays. Combining chromatin interaction data and expression quantitative trait loci, we nominated genes targeted by candidate CREs and prioritized effector genes for AOA and COA.

**Results:** Heritability enrichment analysis suggested a shared role of immune cells in the development of both AOA and COA while highlighting the distinct contribution of lung structural cells in COA. Functional fine-mapping uncovered 21 and 67 credible sets for AOA and COA, respectively, with only 16% shared between the two. Notably, one-third of the loci contained multiple credible sets. Our CRE prioritization strategy nominated 62 and 169 candidate CREs for AOA and COA, respectively. Over 60% of these candidate CREs showed open chromatin in multiple cell lineages, suggesting their potential pleiotropic effects in different cell types. Furthermore, COA candidate CREs were enriched for enhancers experimentally validated by MPRA in BECs. The prioritized effector genes included many genes involved in immune and inflammatory responses. Notably, multiple genes, including *TNFSF4*, a drug target undergoing clinical trials, were supported by two independent GWAS signals, indicating widespread allelic heterogeneity. Four out of six selected candidate CREs demonstrated allele-specific regulatory properties in luciferase assays in BECs.

**Conclusions:** We present a comprehensive characterization of causal variants, regulatory elements, and effector genes underlying AOA and COA genetics. Our results supported a distinct genetic basis between AOA and COA and highlighted regulatory complexity at many GWAS loci marked by both extensive pleiotropy and allelic heterogeneity.

## Background

Asthma is a common, complex lung disease with a significant genetic component [1–3]. The largest genome-wide association study (GWAS) of asthma to date leveraged data from more than 1.5 million individuals across multiple ancestries and identified more than 150 significant loci [4]. Genetic associations, however, do not reveal the molecular mechanisms underlying the pathogenesis of asthma. Moreover, while most people with a diagnosis of asthma share a similar set of symptoms, there are many subtypes in which individuals display unique sets of clinical characteristics and biomarker measurements [5–13]. Age of onset is an important criteria used in differentiating asthma subtypes [14], and recent GWAS of adult-onset asthma (AOA) and childhood-onset asthma (COA) suggested that the clinical heterogeneity between AOA and COA reflected differences in their underlying genetics [15,16].

Yet, challenges remain in translating GWAS results into biological insights [17,18]. Causal variants at individual loci are often elusive due to complex linkage disequilibrium (LD) structures. Furthermore, most associated variants map within non-coding regions of the genome, making it difficult to know their functional effects. It is generally assumed that non-coding GWAS variants exert their effects through changes in the expression of nearby genes. However, identifying the causal genes targeted by GWAS variants is not straightforward due to complexities of gene regulation, with *cis*-regulatory elements (CREs) often regulating more than one gene in more than one cell type or tissue and often over long distances [19,20]. Efforts have been made to address these post-GWAS challenges by jointly analyzing GWAS results with expression quantitative trait loci (eQTLs), using techniques such as colocalization [21–25] and transcriptome-wide association studies [24,26–29] (TWAS). Nonetheless, eQTLs only explain a small fraction of the heritability for most complex traits [30,31], limiting their utility for identifying effector genes. Some studies have explored additional molecular phenotypes such as alternative splicing (s)QTLs [32], chromatin accessibility (ca)QTLs [33,34], and DNA methylation (me)QTLs [35], but these QTLs are not widely available and some (caQTLs and meQTLs) do not point to their target genes directly. Importantly, colocalization and TWAS are prone to false positive findings [36–38]. In addition, to our knowledge, no studies to date have systematically explored the genetic underpinnings of AOA and COA in fine-mapping studies or by integrating functional data from a diverse range of cell types and modalities.

In this study, we used a combination of computational and experimental approaches to systematically fine-map genetic loci associated with AOA and COA [15]. Our innovative pipeline identified putative causal variants at these loci, nominated and validated candidate CREs that are likely disrupted by putative causal variants, and prioritized effector genes supported by multiple lines of genetic evidence (**Fig. 1**). Collectively, our analyses highlighted distinct genetic bases of AOA and COA while revealing pervasive pleiotropy and allelic heterogeneity at asthma GWAS loci.

**Fig. 1.**
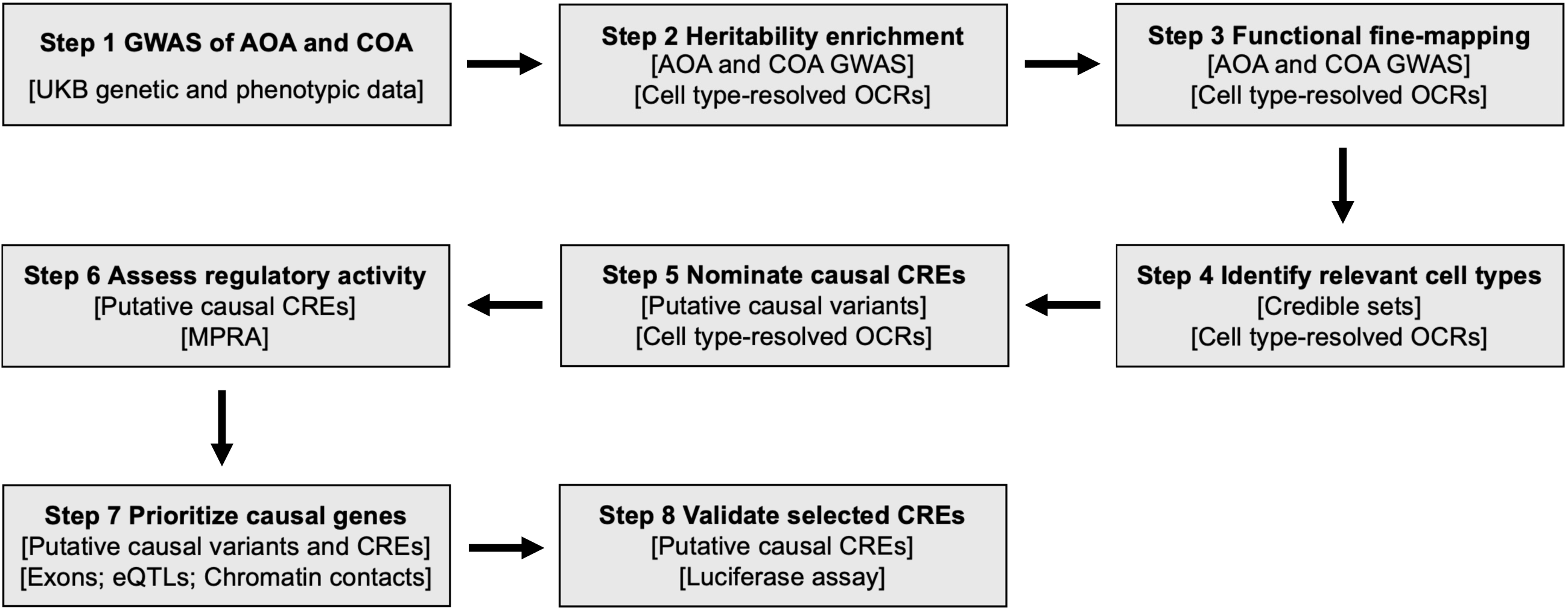
Study workflow. For each step, the input data and assay are shown in brackets. UKB: UK Biobank, OCR: open chromatin region, CRE: *cis*-regulatory element, MPRA: massively parallel reporter assay, eQTL: expression quantitative trait locus.

## Methods

### GWAS

We conducted GWAS of AOA and COA using an updated version of the UKB genotypes following the same protocol and using the same definition as in our previous study [15] (**Additional file 1: Supplementary Methods**; **Additional file 2: Table S1**).

### Heritability enrichment analysis

We performed ATAC-seq in airway smooth muscle cells (ASMCs) and harmonized chromatin accessibility data in 19 lung and seven blood cell types from three published studies [39–41] (**Additional file 1: Supplementary Methods**; **Additional file 2: Table S2**). Using stratified LD score regression [42] (S-LDSC), we estimated the heritability enrichment of open chromatin regions (OCRs) in individual cell types and cell lineages. We adjusted for annotations included in the baseline LD model [43] in all S-LDSC analyses.

### Statistical fine-mapping

Functional fine-mapping was implemented through a two-step procedure. In the first step, we used an empirical Bayesian model, TORUS [44], to estimate a prior probability (functional prior) for each SNP using GWAS summary statistics and a set of input functional annotations (i.e., OCRs in cell lineages significantly enriched for AOA/COA heritability). Next, we used the summary statistics version of “sum of single effects” (SuSiE) [45,46] model to fine-map LD blocks at GWAS loci. For each SNP, SuSiE estimates a posterior inclusion probability (PIP), which reflects the strength of evidence supporting it as a causal variant. To integrate functional annotations with fine-mapping, we specified the prior probabilities of individual SNPs using the functional priors computed by TORUS in the first step. For comparison, we also performed fine-mapping without using functional information (i.e., using uniform priors). See **Additional file 1: Supplementary Methods** for details.

We considered AOA and COA credible sets at the same LD block as shared if they shared more than half of their SNPs or if the total PIP of the shared SNPs was greater than 50% of the PIP of either credible set. If these criteria were not met, the credible set was considered to be AOA- or COA-specific.

### Mapping candidate CREs and computing element PIPs (ePIPs)

To map candidate CREs, we merged the ATAC-seq peaks across 20 lung and seven blood cell types [39,40] using the bedtools [47] version 2.30.0 `merge` command with the `-d` option set to -1. The output was a set of non-overlapping OCRs, each of which was considered as a candidate CRE. The ePIP of a candidate CRE was defined as the sum of the PIPs of the credible set SNPs within the CRE.

### Linking candidate CREs to target genes

We used four features to nominate likely target genes for each candidate CRE: 1) the nearest gene to the candidate CRE; 2) if the SNP with the highest PIP in the candidate CRE was an eQTL [21,48–51] for that gene; 3) if the candidate CRE interacted with the promoter of a gene by Promoter Capture Hi-C (PCHi-C) [41,52,53], defined as ≥ 50% physical overlap between the candidate CRE and the non-promoter end of PCHi-C loops; and 4) if a regulatory element in the activity-by-contact (ABC) dataset [54] had ≥ 50% physical overlap with the candidate CRE, in which case the gene with the highest ABC score was considered as a putative target gene. For each candidate CRE, we only used eQTL, PCHi-C, and ABC data collected from tissues and/or cell types matching the CRE’s cell lineage(s) to identify likely target genes (**Additional file 2: Table S2**). Gene annotations were curated using mapgen R package [55] version 0.5.9, and we restricted our analysis to protein-coding genes.

### Prioritizing effector genes

To prioritize candidate genes, we utilized both candidate CREs and exonic variants, which may disrupt protein coding sequences. The derived gene scores summarized the total genetic evidence supporting the role of a gene in AOA or COA. The score of a gene 𝑔, 𝑆_!_, was the sum of the contributions of all variants that support 𝑔. We defined genes with gene score ≥ 0.95 as high-confidence candidate causal genes for AOA or COA. See **Additional file 1: Supplementary Methods** for details. To attribute a gene’s score to individual credible sets, we grouped the variants linked to the gene by credible sets and calculated the total contribution of each credible set to the gene.

## Results

### Leveraging functional annotations to fine-map AOA and COA GWAS loci

To identify causal variants of asthma, we statistically fine-mapped AOA and COA loci from GWAS in UKB (**Methods**; **Additional file 1: Fig. S1**). We employed a functional fine-mapping approach, which leveraged chromatin accessibility data from blood and lung cell types relevant to asthma pathogenesis to improve the resolution of fine-mapping. Using ATAC-seq peaks (**Additional file 2: Table S2**), we first mapped OCRs of each cell type. Because OCRs in cell types from the same cell lineage shared similar heritability enrichments (**Additional file 1: Fig. S2**), we pooled OCRs by lineage for assessing heritability enrichment and for all subsequent analyses (**Methods**). Both AOA (p = 6.98 × 10^-5^) and COA (p = 6.13 × 10^-9^) risk variants were significantly enriched in OCRs of lymphocytes (**Fig. 2A**). COA risk variants were also significantly enriched in OCRs of epithelial cells (p = 0.02) and mesenchymal cells (p = 0.02).

**Fig. 2.**
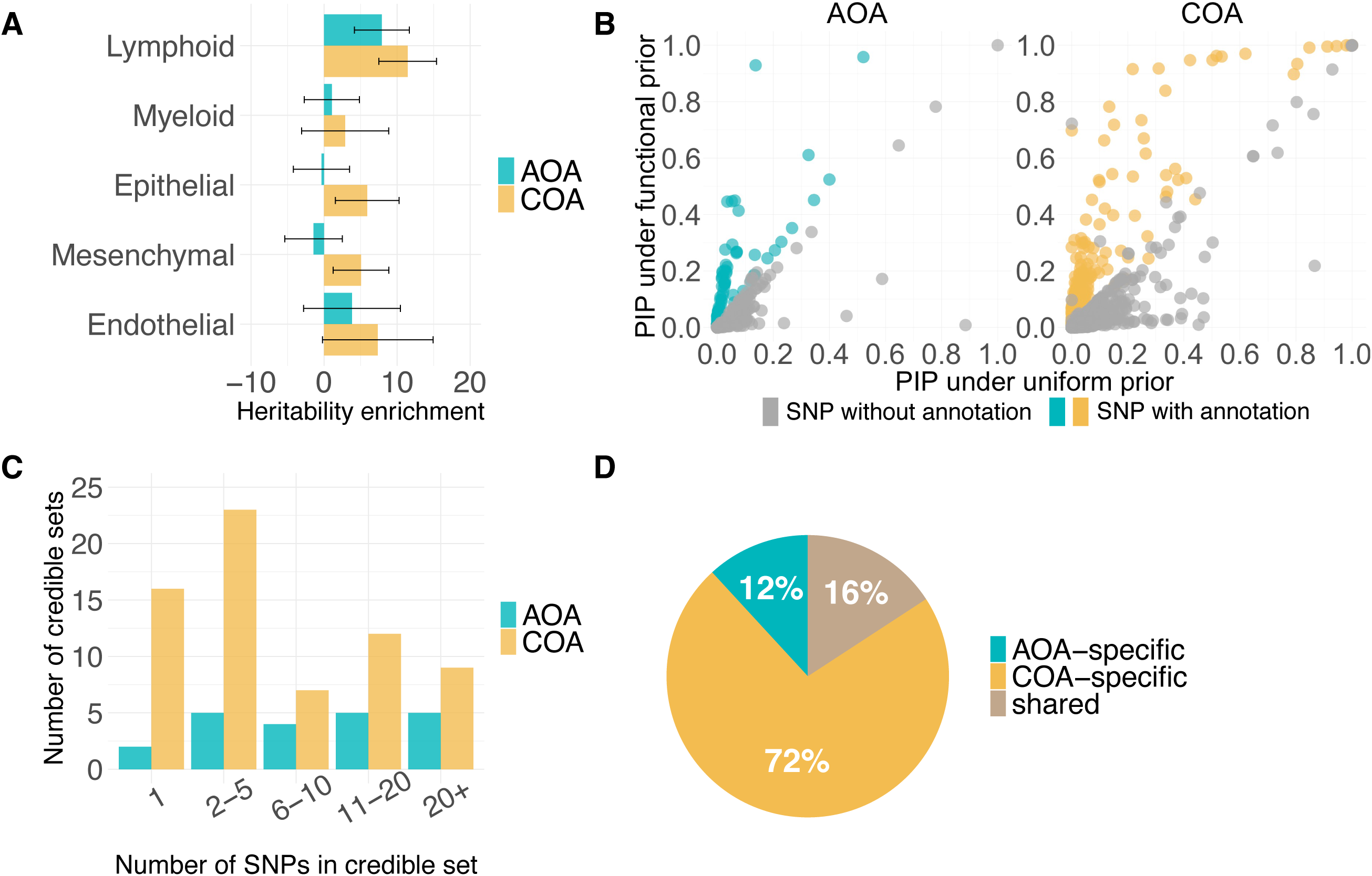
A. Heritability enrichment estimates for OCRs in asthma-relevant cell types. Lymphoid: lung B cells, lung T cells, lung NK cells, blood B cells, blood T cells, blood NK cells; Myeloid: lung macrophage, blood myeloid dendritic cells, blood plasmacytoid dendritic cells, blood monocytes; Epithelial: alveolar type 1 cells, alveolar type 2 cells, pulmonary neuroendocrine cells, lung basal cells, lung ciliated cells, lung club cells, BECs; Mesenchymal: lung matrix fibroblasts, lung myofibroblasts, lung pericytes, ASMCs; Endothelial: lung arterial cells, lung capillary cells, lung lymphatic cells. Confidence intervals are ± 2 standard errors. **B.** PIPs for SNPs in adult-onset asthma (left panel) and childhood-onset asthma (right panel) fine-mapping, with SNPs weighted by functional annotations (y-axis) or by uniform weights (x-axis). **C.** Distribution of the number of SNPs in the adult-onset asthma and childhood-onset asthma credible sets. **D.** Distribution of the number of shared and specific credible sets.

Next, we integrated OCRs of enriched cell lineages using a Bayesian hierarchical model [44] (**Additional file 2: Table S3**) and performed functional fine-mapping [45,46] for all LD blocks harboring at least one genome-wide significant SNP (p < 5 × 10^-8^) (**Methods**; **Additional file 1: Supplementary Methods**). For comparison, we also performed fine-mapping [45,46] under the default setting that assumes all SNPs are equally likely to be causal a priori (**Fig. 2B**). To quantify the effect of incorporating functional information in fine-mapping, we assigned variants into one of three categories by their PIP: low-confidence (0.1 < PIP ≤ 0.5), mid-confidence (0.5 < PIP ≤ 0.8), and high-confidence (PIP > 0.8) (**Additional file 2: Table S4**). For AOA, functional fine-mapping led to a 30% increase in low-confidence variants, no change in mid-confidence variants, and a 50% increase in high-confidence variants. For COA, we observed 7%, 120%, and 33% increase in low-confidence, mid-confidence, and high-confidence variants, respectively.

Fine-mapping identifies groups of variants (i.e., credible sets) that contain at least one causal variant with 95% confidence. We discovered 21 and 67 credible sets among the 16 and 48 LD blocks that were fine-mapped for AOA and COA, respectively (**Additional file 2: Table S5**). About one-third of the LD blocks (5 AOA and 16 COA) had more than one credible set (**Additional file 1: Fig. S3**), suggesting multiple independent causal signals within these blocks. The number of SNPs within a credible set varied widely, ranging from 1 to 136, with median values of 10 for AOA and 4 for COA (**Fig. 2C**). Among all credible sets, only 16% were shared between AOA and COA (**Fig. 2D**).

### Identifying cell types and CREs mediating genetic risk of asthma

The enrichment analysis above revealed genome-wide cell type heritability enrichment patterns but did not provide information on the relevant cell types for individual credible sets. To assess the evidence supporting a cell lineage for each credible set, we calculated the proportion of PIPs of variants that were within OCRs of each lineage (**Additional file 1: Supplementary Methods**). This proportion can be understood as the probability that the causal variant acts on the phenotype through a lineage. Using this strategy, we found that among AOA credible sets, 75% of the PIPs on average were attributed to lymphocytes (**Fig. 3A**). In contrast, the COA credible sets had higher proportions of PIPs attributed to epithelial cells (19%) and mesenchymal cells (17%), in addition to lymphocytes (46%) (**Fig. 3B**).

**Fig. 3.**
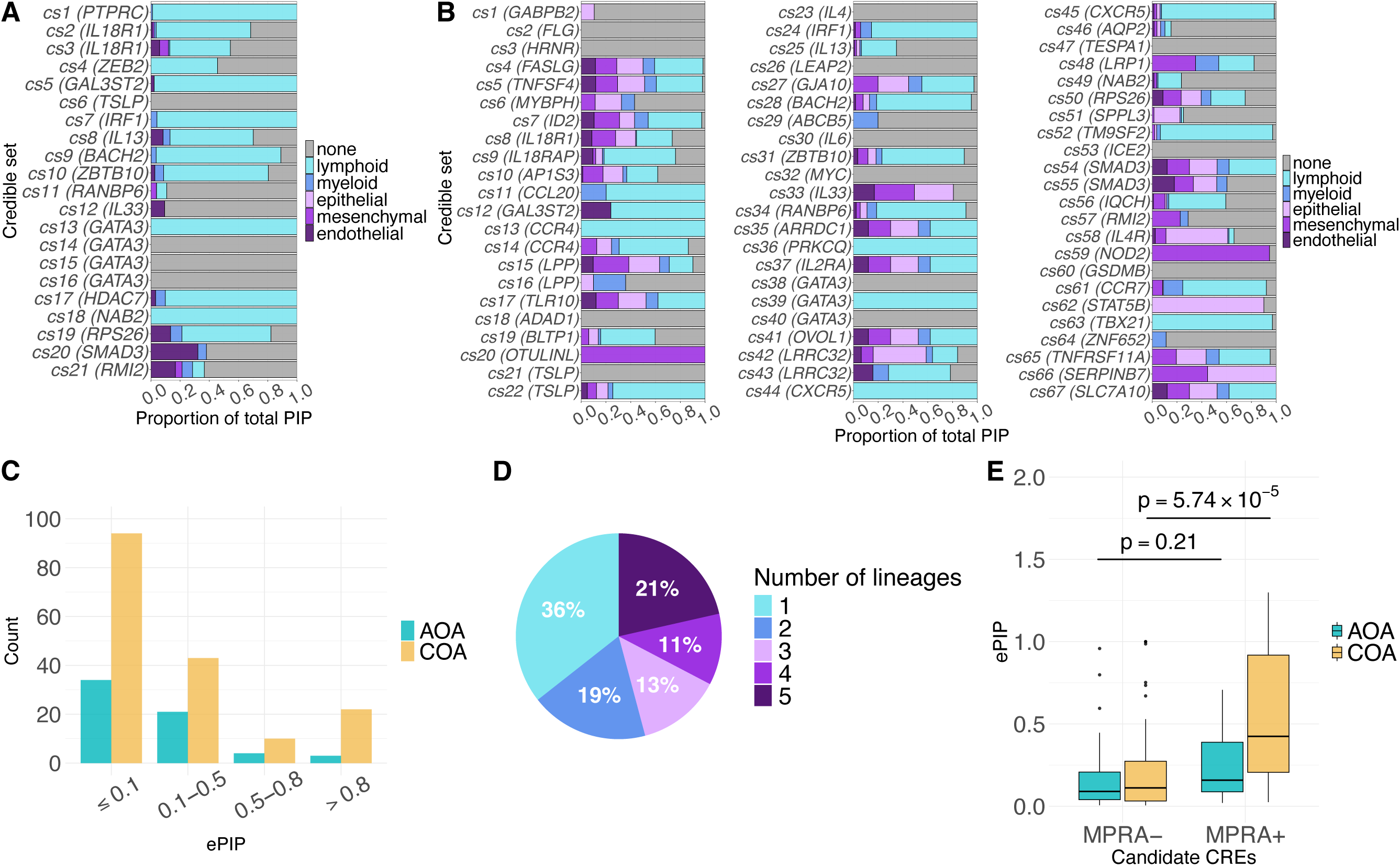
**A.** Cellular contexts of adult-onset asthma credible sets based on OCRs. The proportion of the total PIP in each credible set is attributed to OCRs of each of the five cell lineages or to none. Each horizontal bar corresponds to a credible set, which is labelled in parentheses by the nearest gene to the SNP with the highest PIP; the length of bars of different colors shows the proportion of PIPs assigned to each lineage. Because not all SNPs in the credible sets overlapped with an OCR, some were not assigned to a cell lineage (gray bars). **B.** Cellular context of childhood-onset asthma credible sets. **C.** Adult-onset asthma and childhood-onset asthma candidate CRE ePIP distributions. **D.** Distribution of the number of cell lineages underlying candidate CREs. **E.** Adult-onset asthma and childhood-onset asthma ePIP distributions of candidate CREs (from panels C and D) that overlapped with bronchial epithelial cells MPRA^+^ and MPRA^-^ sequences. The p-values were computed using Wilcoxon rank-sum test.

We next sought to nominate specific CREs that may be mediating the genetic effects of causal variants. We ranked candidate CREs by their ePIPs, which can be interpreted as the expected number of causal SNPs targeting the CRE (**Methods**). Using this approach, we identified 62 AOA and 169 COA candidate CREs with nonzero ePIPs (**Fig. 3C; Additional file 2: Table S6**). Notably, 64% of these candidate CREs were defined by OCRs of multiple cell lineages (**Fig. 3D**), indicating potential pleiotropic effect of asthma risk variants.

To experimentally assess regulatory activities of candidate CREs and variants, we performed MPRA in a human bronchial epithelial cell (BEC) line, 16HBE14o-, to examine enhancer activities and allele-specific effects of 2,034 SNPs chosen from AOA and COA GWAS loci (**Additional file 1: Supplementary Methods**). Among those, 438 SNPs were in sequences that tested positive for enhancer activity in a bronchial epithelial cell line, and 34 of those showed allele-specific effect on enhancer activity (**Additional file 2: Table S7**). We then used the MPRA results to assess the CREs selected by our ePIP strategy (**Additional file 1: Supplementary Methods**). The validated sequences in MPRA (MPRA^+^ set) had significantly higher ePIPs than sequences tested negative in MPRA (MPRA^-^ set) for COA (p = 5.74 × 10^-5^, **Fig. 3E**). In contrast, we did not observe a significant difference in ePIPs for AOA (p = 0.21, **Fig. 3E**). Taken together, the MPRA results suggest that the COA candidate CREs were distinctly enriched for enhancer activities in BECs.

### Linking candidate CREs to target genes and prioritizing asthma risk genes

We next aimed to link candidate CREs to their target genes and identify likely causal genes for AOA and COA. Using functional genomics data that capture long-range regulation, including chromatin interactions and eQTLs from asthma-relevant tissues and cell types (**Methods**; **Additional file 2: Table. S2**), we nominated 107 putative target genes for 62 AOA candidate CREs and 253 putative target genes for 169 COA candidate CREs. Notably, 53 and 118 candidate CREs were linked to at least two different genes in AOA and COA, respectively (**Additional file 1: Fig. S4**), thus underscoring the challenge of precisely identifying target genes for specific CREs.

To address this challenge, we developed a scoring strategy to prioritize asthma effector genes by aggregating causal evidence from variants linked to the same gene (**Methods**). Using this strategy, we identified 76 AOA genes and 203 COA genes with nonzero gene score (**Additional file 2: Table S8**). Of these, 10 and 35 genes were considered as high-confidence candidate causal genes for AOA and COA, respectively (**Fig. 4A**, **Fig. 4B**). The most significant genes were often supported by multiple lines of evidence, with the greatest contributions coming from variants potentially targeting their nearest genes and variants linked to the genes by ABC models. All AOA candidate causal genes were targeted by SNPs from a single credible set, while 11 COA candidate causal genes were supported by SNPs from more than one credible set (**Additional file 1: Fig. S5**).

**Fig. 4.**
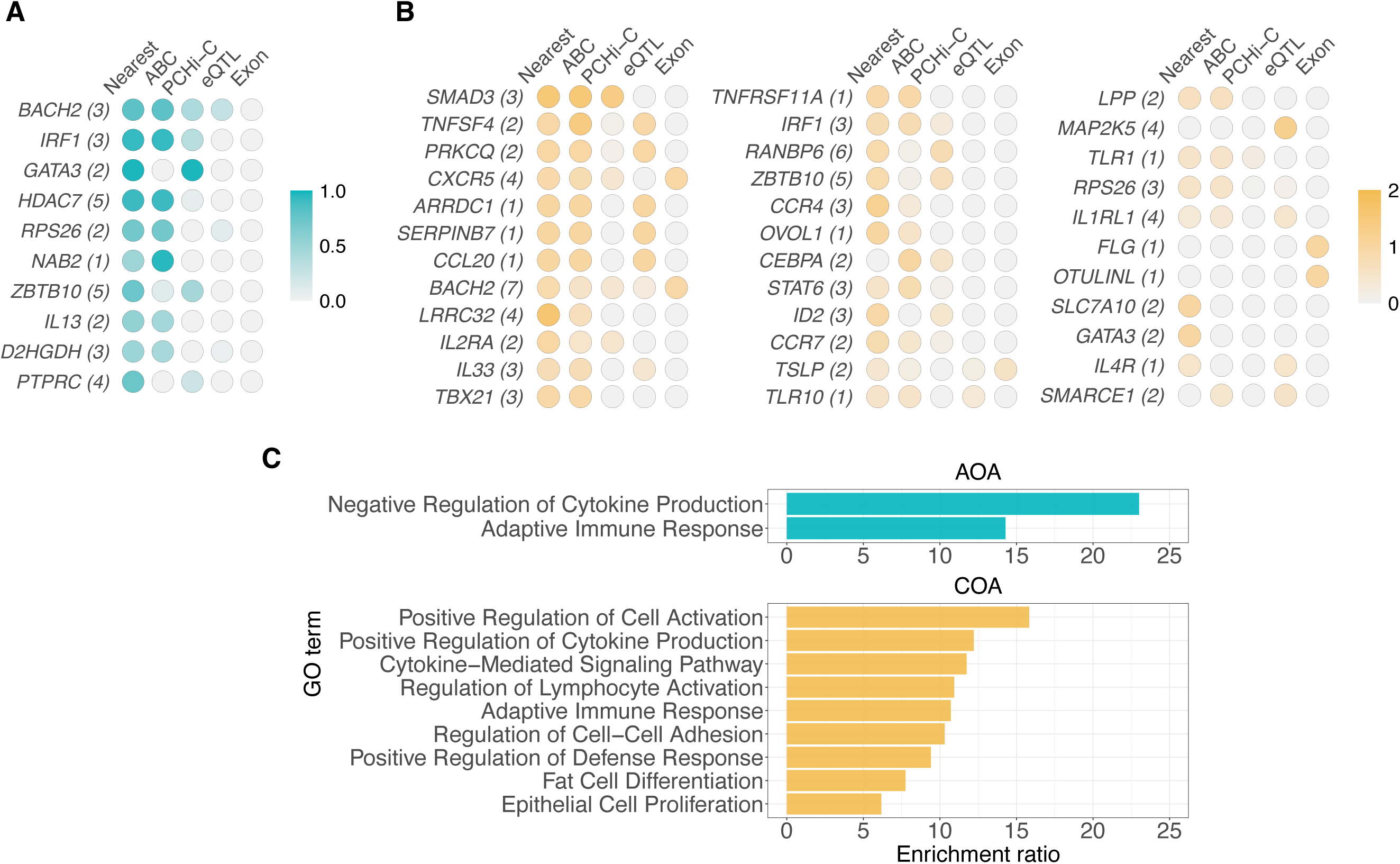
**A.** Adult-onset asthma high-confidence candidate causal genes (N = 10), listed in descending order. The intensity of color shows the score contributed by each category. Nearest: variants whose nearest gene is the candidate gene; ABC: variants linked to the candidate gene by the ABC model; PCHi-C: variants linked to the candidate gene by PCHi-C; eQTL: variants linked to the candidate gene by eQTL; Exon: variants in the candidate gene’s exonic regions. The number in the parentheses indicates the number of variants linked to the corresponding gene. **B.** Childhood-onset asthma high-confidence candidate causal genes (N = 35), listed in descending order. **C.** Top Biological Processes GO terms enriched among AOA (top) and COA (bottom) high-confidence candidate causal genes, generated by WebGestalt’s weighted set cover algorithm.

To understand the biological functions of the prioritized genes, we assessed the enrichment of Biological Process GO terms [56,57] for candidate risk genes (**Additional file 1: Supplementary Methods**). A total of nine and 56 Biological Process GO terms were significantly enriched for AOA and COA candidate genes (FDR < 0.05), respectively, with eight of these shared between AOA and COA (**Additional file 2: Table S9**). The top enriched GO terms in both AOA and COA were associated with cytokine production and inflammatory response (**Fig. 4C**).

### Functionally assaying candidate CREs and causal variants

Based on our integrative analyses, we selected six candidate CREs at both shared and specific loci for further functional validation using luciferase assays in 16HBE14o-cells (**Additional file 1: Supplementary Methods**): three were high-confidence candidate enhancers, two were candidate enhancers whose top SNP overlapped with an MPRA^+^ sequence, and one was in a promoter region (**Additional file 2: Table S10**). The results are described in the following sections.

#### Candidate enhancers at a COA-specific locus at chromosome 1q25.1

We identified one credible set containing two SNPs at a COA-specific locus at chromosome 1q25.1 (**Fig. 5A**; **Additional file 2: Table S5**, COA cs5). The most significant SNP in COA GWAS and the most likely causal SNP, rs11811856 (PIP = 0.95), mapped within an intron of *TNFSF4* and overlapped with an OCR in epithelial, endothelial, mesenchymal, and myeloid cells in lung and lymphocytes in blood. The distance from the OCR midpoint to the *TNFSF4* transcription start site (TSS) was 4,966 bp. This OCR physically contacted the promoters of several genes based on PCHi-C of blood immune cells [52]: *TNFSF18*, *CENPL*, and *DARS2*. To complement the PCHi-C results, we checked ABC scores in relevant cell types (**Additional file 2: Table S2**).

**Fig. 5.**
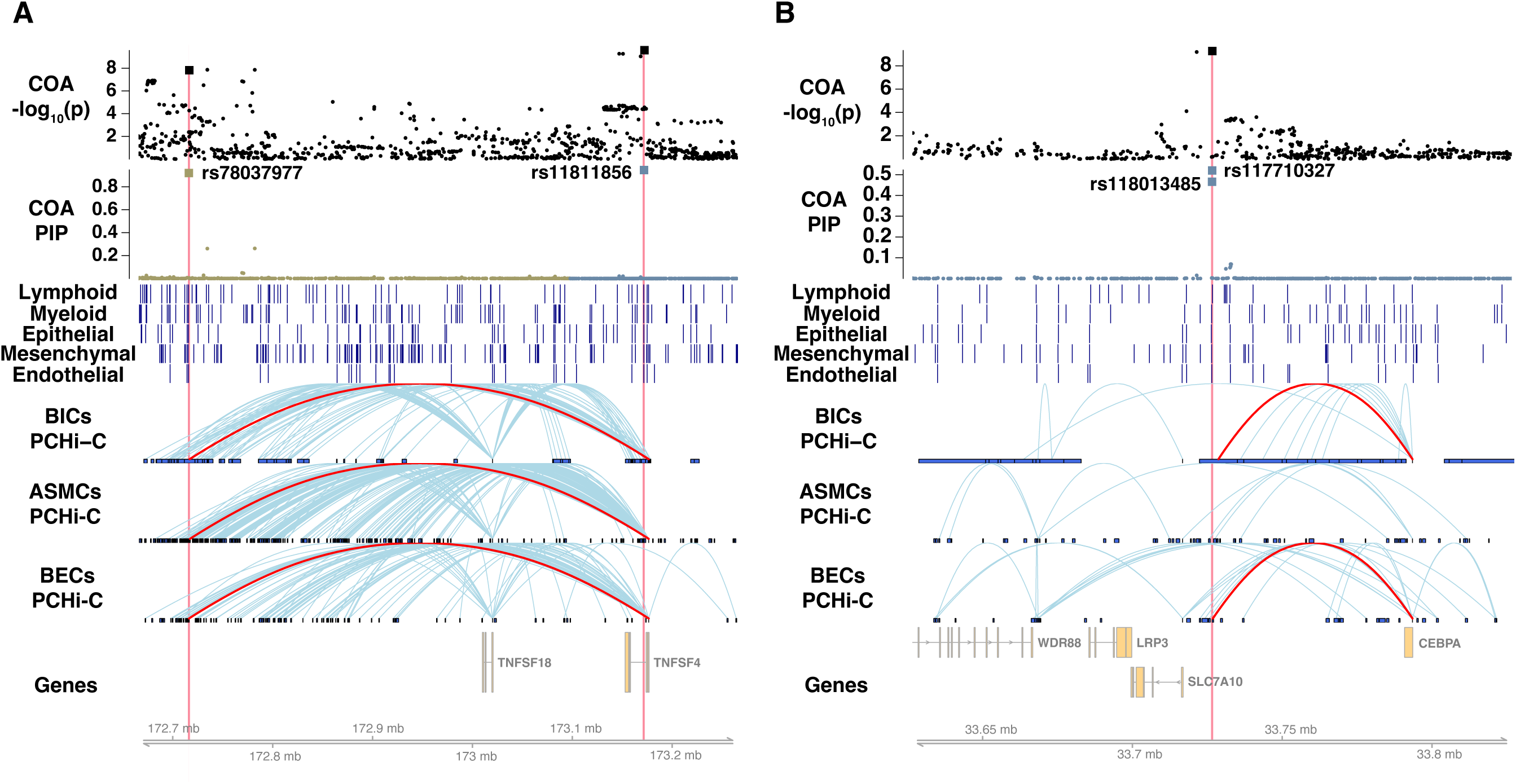
**A.** A childhood-onset asthma-specific locus at chromosome 1q25.1. From top to bottom, the first two tracks show the -log10 p-values from GWAS and PIPs from fine-mapping, respectively. Each point is a SNP, and assayed SNPs are denoted by larger squares. Different colors are used in the PIP track to represent different LD blocks. The two SNPs in candidate enhancers, rs78037977 and rs11811856, are highlighted in red. The next five tracks display chromatin accessibility from (sn)ATAC-seq of different cell lineages, with each dark blue vertical bar showing the location of an OCR. The next three tracks show chromatin interactions from PCHi-C of different cells, where the loops from the distal candidate enhancer to *TNFSF4* promoter in all three cell types are highlighted in red. The last track shows the genes at the locus. BICs: blood immune cells, ASMCs: airway smooth muscle cells, BECs: bronchial epithelial cells. **B.** A COA-specific locus at chromosome 19q13.11. The PCHi-C loops from the candidate enhancer to the *CEBPA* promoter in blood cells and bronchial epithelial cells are highlighted in red.

Interestingly, *TNFSF4* is the most likely target of the OCR based on the ABC scores in immune cells, fibroblasts, and endothelial cells [54]. We also identified a distal candidate enhancer that looped to the promoter of *TNFSF4* in PCHi-C of blood immune cells, BECs, and ASMCs (distance = 460,760 bp) harboring a high-PIP SNP rs78037977 (PIP = 0.92; **Additional file 2: Table S5**, COA cs4). This OCR overlapped with an MPRA^+^ sequence, supporting its regulatory activity. While this enhancer also contacted dozens of other gene promoters according to PCHi-C, *TNFSF4* was a top gene by ABC scores in immune cells, epithelial cells, and fibroblasts. Taken together, these observations suggested that *TNFSF4* is likely a COA risk gene, possibly with two independent causal signals targeting this gene.

Luciferase assay showed enhancer activity only for the rs11811856-C allele, the non-risk allele (**Additional file 1: Fig. S6**, left), for the candidate enhancer in the *TNFSF4* intron, suggesting allele-specific effects of rs11811856-C vs. rs11811856-G (p = 0.02). In contrast, the distal candidate enhancer did not show regulatory effect in the luciferase assay (**Additional file 1: Fig. S6**, right), possibly due to cell type-specific regulation in a different cell type.

#### A candidate enhancer at a COA-specific locus at chromosome 19q13.11

One credible set (**Additional file 2: Table S5**, COA cs67) at a COA-specific locus at chromosome 19q13.11 (**Fig. 5B**) had two putative causal SNPs, rs118013485 (PIP = 0.46) and rs117710327 (PIP = 0.52). The two SNPs were in nearly perfect LD, with only two haplotypes (rs118013485-G/rs117710327-C and rs118013485-A /rs117710327-A) observed in the 1000 Genomes European populations [58,59]. Both SNPs resided in the same OCRs in cell types from all five lineages. The ePIP of this candidate sequence was 0.99, suggesting that it mostly likely mediates the effect of the underlying causal variant(s). Although none of the eQTL datasets identified any target gene(s) for this candidate enhancer, PCHi-C in BECs [41] indicated that this candidate CRE only contacted the promoter of *CEBPA* (distance = 66,853 bp). In blood immune cells, this candidate enhancer looped to the promoters of *CEBPG* and *CEBPA*, both of which are CCAAT enhancer binding proteins. Moreover, *CEBPA* had the highest ABC score in immune cells. We observed different levels of enhancer activities (rs118013485-A/rs117710327-A vs. rs118013485-G/rs117710327-C, p = 0.04) in luciferase assays between the two haplotypes (**Additional file 1: Fig. S7**), with decreased activity associated with the haplotype carrying the COA risk alleles.

#### Candidate enhancers at an AOA and COA shared locus at 5q31.1

At a shared locus at 5q31.1, we discovered two credible sets shared by AOA and COA (**Additional file 2: Table S5**, AOA cs7 and COA cs24, AOA cs8 and COA cs25), and one credible set that was specific to COA (**Additional file 2: Table S5**, COA cs23). Nominating the true causal SNPs from these credible sets was difficult, as none of the SNPs in the five credible sets had a PIP > 0.5. Therefore, we selected the AOA credible set containing the fewest SNPs for functional validation studies (AOA cs7; **Fig. 6A**). Among the four SNPs in cs7, rs1023518 (PIP = 0.35) and rs3857440 (PIP = 0.30) were captured by one candidate enhancer, while SNP rs3749833 (PIP = 0.27) resided in a separate candidate enhancer, both of which were represented by OCRs in all five blood and cell lineages. These three SNPs together accounted for 97% of the total PIP in the credible set. Furthermore, rs1023518 and rs3749833 overlapped with different sequences that each demonstrated enhancer activity in MPRA, whereas the sequence containing rs3857440 did not show enhancer activity in MPRA.

**Fig. 6.**
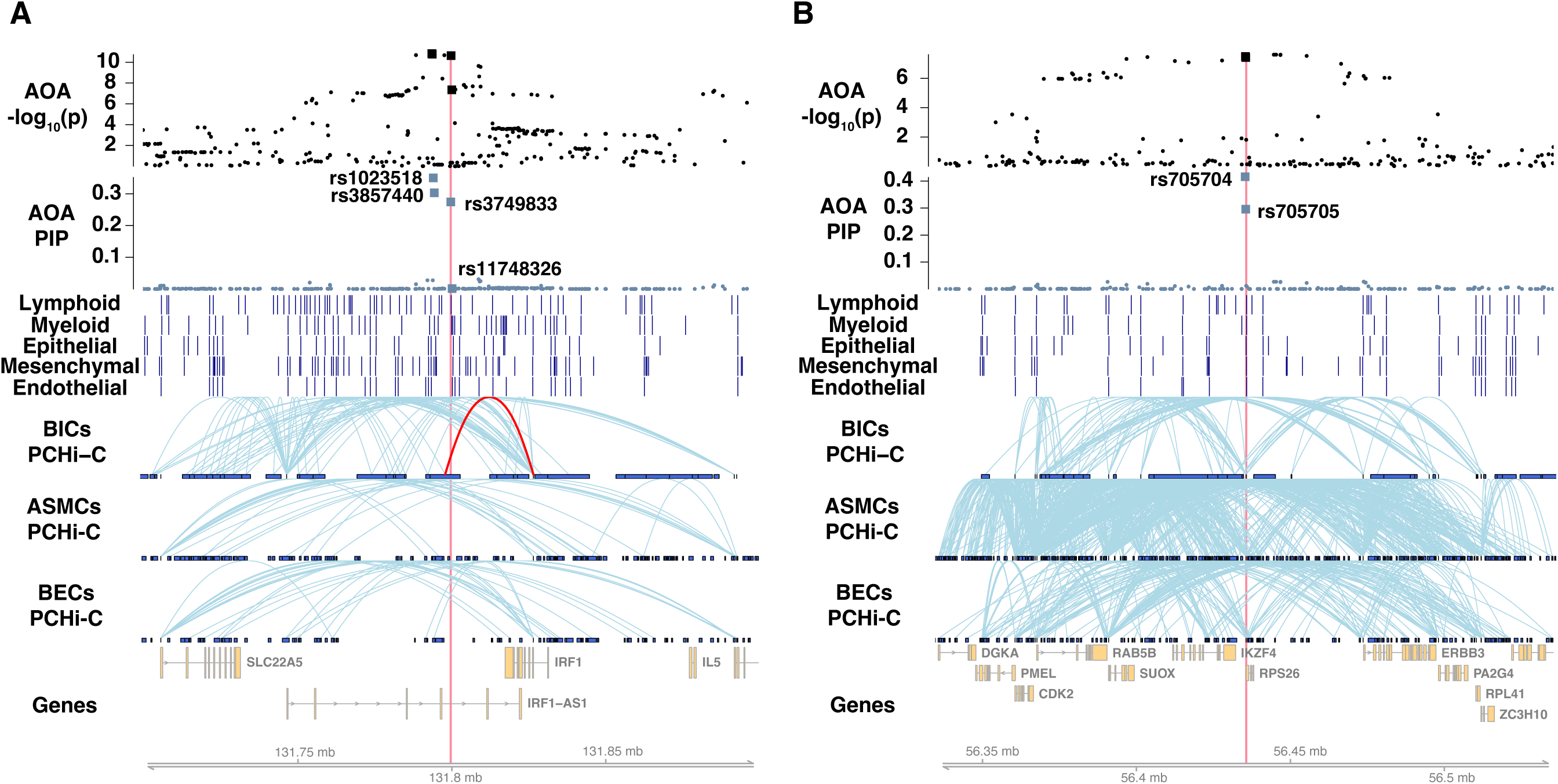
**A.** A shared locus at chromosome 5q31.1. See Fig. 5 figure legend. The PCHi-C loop from the candidate enhancer to *IRF1* promoter is highlighted in red in blood immune cells. **B.** A shared locus at chromosome 12q13.2.

In luciferase assays, the sequence harboring rs1023518 and rs3857440 tested negative (**Additional file 1: Fig. S8**, left), but the sequence containing rs3749833 was validated as an allele-specific enhancer (**Additional file 1: Fig. S8**, right). Moreover, although another genome-wide significant SNP rs11748326 was located within the same luciferase construct as rs3749833, only rs3749833 showed significant allelic effects (**Additional file 1: Fig. S8**, right). These observations indicated that rs3749833 is likely a causal variant exerting its effect in BECs. We also looked at eQTLs and chromatin interaction data to determine the likely target genes of this enhancer. While the GTEx eQTL data nominated *PDLIM4* as the putative target gene in skin and *SLC22A5* as the putative target gene in whole blood, lung, and skin, this enhancer only interacted with the promoter of *IRF1* in PCHi-C of blood immune cells. Additionally, *IRF1* had the highest ABC score in immune cells. These findings suggest that one or more of these genes are regulated by this enhancer.

#### A candidate promoter at an AOA and COA shared locus at 12q13.2

We evaluated an OCR located 2 kb upstream of *RPS26* that was characterized by ATAC-seq peaks in all 27 blood and lung cell types. The AOA and COA ePIPs of this OCR were 0.71 and 0.51, respectively, attributed to two SNPs in a pair of shared credible sets: rs705704 (AOA PIP = 0.41; COA PIP = 0.38) and rs705705 (AOA PIP = 0.29; COA PIP = 0.13) (**Additional file 2: Table S5**, AOA cs19 and COA cs50). We observed extensive chromatin interactions at this locus in BECs and ASMCs, potentially indicating a high level of regulatory activities in these cell types (**Fig. 6B**).

We performed luciferase assays for the two haplotypes comprised of the two SNPs in 1000 Genomes European populations (rs705704-G/rs705705-G and rs705704-A/rs705705-C). In line with our expectations for a promoter, we observed strong regulatory effect of both haplotypes on the luciferase activity, with fold change compared to the control ranging from ∼50 times to > 300 times across experimental replicates (**Additional file 1: Fig. S9**). In addition, the asthma-associated rs705704-A/rs705705-C haplotype showed significantly lower luciferase activity than the rs705704-G/rs705705-G haplotype (rs705704-A/rs705705-C vs. rs705704-G/rs705705-G, p = 0.01), suggesting haplotype-specific regulation.

## Discussion

Personalized risk prediction and treatment strategies for common, complex diseases are the aspirations of precision medicine [60]. The extraordinary heterogeneity of asthma makes these goals particularly challenging. Having a more refined understanding of the shared and distinct molecular genetic mechanisms underlying different asthma subtypes could lead to the discovery of new therapeutic targets as well as identifying individuals who would most likely benefit from therapies. Indeed, a recent study [61] estimated that drugs targeting genes with genetic support had 2.6 times greater probability of achieving clinical success than those targeting genes without genetic support. Our GWAS of AOA and COA [15] serves as a first step toward characterizing the underlying molecular mechanisms and nominating causal genes for these two important asthma subtypes.

In this study, we coupled computational and experimental methods to systematically uncover putative causal variants, candidate CREs, and effector genes for AOA and COA. Utilizing chromatin accessibility data in multiple cell types, we showed that both AOA and COA GWAS signals were concentrated in regulatory regions of immune cells. In contrast, the enrichment of GWAS loci in lung structural cells was a distinctive feature of COA, consistent with results in our previous GWAS [15] based on gene expression data. Leveraging functional fine-mapping, we uncovered a plethora of causal signals that further highlighted the distinct genetic bases underlying risk for AOA and COA. Our multi-level analyses revealed two broad patterns: first, most candidate CREs of asthma were in OCRs across multiple cell lineages, suggesting that most genetic variants of asthma have pleiotropic effects across cell types. Second, allelic heterogeneity was common. This was supported by both the presence of more than one credible set at many loci, and by the fact that many of the candidate genes were supported by two independent causal signals. Overall, our results underscore the complexity of the molecular mechanisms linking genetic variants of asthma pathogenesis.

Many of the genes prioritized by our scoring system had strong prior evidence supporting their roles in asthma. To highlight a few examples: (1) *BACH2*, the highest-ranked AOA risk gene, is a key regulator of T-cell and B-cell differentiation [62–64]. (2) *SMAD3*, the highest-scoring COA effector gene, is a crucial transcription factor in the transforming growth factor beta signaling pathway [65], a central mediator of airway remodeling in asthma [66]. (3) *GATA3*, the third highest-scoring AOA effector gene and a COA candidate risk gene, is a master regulator [67,68] that modulates the expression and production of the type 2 cytokines that play a prominent role in both AOA and COA [69,70]. (4) The prioritized causal genes of COA included several epithelial function-related genes. *OVOL1*, a transcription factor involved in epithelial cell differentiation [71], regulates the expression of *FLG* in normal human epidermal keratinocytes [72].

The *OVOL1*-*FLG* axis contributes to the pathogenesis of atopic dermatitis, an allergic condition that is often comorbid with asthma and has shared genetics with asthma [73], likely mediated by disrupting barrier function [74]. (5) The two prioritized toll-like receptor (TLRs) genes, *TLR1* and *TLR10*, are both expressed in airway epithelium [75]. Together with other TLRs, they orchestrate response against microbes through the activation of TLR signaling pathways in epithelial cells [76,77]. (6) Finally, *HDAC7*, the fifth highest scoring gene for AOA, resides at an AOA-specific GWAS locus. *HDAC7* is a histone deacetylase involved in transcriptional regulation. This gene plays a key role in the function of regulatory T cells [78] and has been shown to potentially play a role in asthma and allergic diseases through epigenetic modifications [79].

The effectiveness of our analytical strategy was supported by the successful validation of selected candidate CREs by luciferase assays: four out of the six selected candidate CREs displayed regulatory activity and allelic effects *in vitro*. The genes likely regulated by these validated CREs were among the top genes prioritized by our gene scores and have known relationships to the pathogenesis of AOA and/or COA. For example, *TNFSF4* encodes OX40 ligand (OX40L). By binding to the OX40 receptor, OX40L on antigen-presenting cells activates the OX40 costimulatory molecule on T cells.

Importantly, the OX40-OX40L pathway has been shown to play a key role in the differentiation of Th2 cells and the activation of memory Th2 cells [80]. A recent Phase 2b clinical trial, STREAM-AD, showed that amlitelimab, a non-T cell depleting monoclonal antibody that blocks OX40L on antigen-presenting cells, exhibited sustained treatment effects on patients with atopic dermatitis [81]. A Phase 2 study examining the efficacy of amlitelimab on asthma patients is underway [81]. Another candidate effector gene, *CEBPA*, is a key regulator of lung epithelial cell development [82,83]. Previous studies showed that CEBPA expression was absent in cultured ASMCs from subjects with asthma [84,85]. The lack of CEBPA expression was associated with the failure of glucocorticoids to inhibit ASMC proliferation [84], suggesting that it could play a role in steroid-resistant asthma. Consistent with this observation, the enhancer potentially regulating *CEBPA* displayed lower activity in luciferase assays in the 16HBE14o-cell line with the risk allele for COA (rs117710327-C) compared to the non-risk rs117710327-A allele. Another putative causal gene, *IRF1*, encodes a transcription factor that regulates the activity of interferon and is involved in various aspects of adaptive and innate immune responses to pathogens [86–88]. *IRF1* is upregulated by rhinovirus (RV) in epithelial cells [89]. RV-associated wheezing illness in early life is one of the most significant risk factors for COA [90,91] and is strongly associated with asthma exacerbations and hospitalizations throughout life [92]. In addition, *IRF1* has been identified as a key driver of lipopolysaccharide (LPS)-induced interferon responses at birth [93]. Based on these and other data [94–96], impaired immune responses to microbial infections is thought to be a key mechanism underlying asthma onset and severity [97]. These findings are also consistent with our observation of reduced enhancer activity associated with the asthma risk allele (rs3749833-C).

We recognize several limitations of our study. First, our analysis primarily relied on open chromatin as functional annotations, missing other mechanisms of gene regulation (e.g., alternative splicing). Second, in regions with extensive LD, many SNPs will receive small PIPs, often making it impossible to distinguish likely causal SNP(s). Both possibilities may explain why the most significant COA locus at chromosome 17q12-q21 was not among those prioritized by our pipeline. This locus is characterized by extensive LD over ∼150 kb in populations of European ancestry and the likely causal SNPs affect splicing (rs11078928) of *GSDMB* (gasdermin B) and/or encode a missense mutation (rs2305480) in *GSDMB* [98]. Third, the epigenomic and gene expression data used in our studies were in unstimulated cells. It is possible that cells stimulated with asthma-promoting cytokines or viruses, as examples, will induce context-specific CREs that were missed by the current study. Fourth, we applied a heuristic method to score and rank the genes and different kinds of evidence linking a CRE with a gene were weighted equally. A better approach may be to assign weights differently, putting more emphasis on datasets more likely to support functional relationships. Fifth, to maximize detection power, we used summary statistics from AOA and COA GWAS for fine-mapping. We therefore were not able to include individual-level comorbidities as covariates in our analyses, nor were we able to stratify our analyses by clinical features. As a result, we may have missed signals that are specific to severe asthma [99] or asthma associated with other disorders [100,101]. Finally, our study included only UKB individuals who self-identified as White British and, therefore, our fine-mapping results reflect the specific LD and allele frequency patterns of this population. A recent study [102] has shown that fine-mapping can greatly benefit from including individuals of diverse genetic backgrounds, which can uncover putative causal variants that are more frequent in non-European populations. Additionally, the distinct haplotype structures among different populations can help disentangle SNPs that are in high LD in one population but not in others. Taken together, increasing the genetic diversity of future fine-mapping studies, along with rigorous analytical approaches and more precise phenotype definitions, is critical for expanding our understanding of the genetic architecture of complex traits across various populations.

## Conclusions

By combining experimental and computational approaches, our study provides the most thorough follow-up of AOA and COA GWAS discoveries to date. We identified numerous risk variants, regulatory elements, and candidate genes and uncovered key insights into the genetic architecture of AOA and COA. Our data sets provide a valuable resource for future functional studies to understand the biological mechanisms underlying the genetics of asthma with onset in both childhood and later in life.

## Availability of data and materials

This study uses genotype and phenotype data from the UK Biobank under application number 44300. Access to UK Biobank resource is available with application at http://www.ukbiobank.ac.uk. Summary statistics of AOA and COA GWAS performed with UK Biobank version 3 genotypes will be made available prior to publication of the manuscript. The sequencing data generated in this study were deposited in EMBL-EBI’s Array Express database (https://www.ebi.ac.uk/biostudies/arrayexpress) under accession numbers E-MTAB-14267 (ASMCs ATAC-seq), E-MTAB-14273 (BECs MPRA), and E-MTAB-14295 (ASMCs PCHi-C). The snATAC-seq data from 18 lung cell types were downloaded from https://www.lungepigenome.org. The ATAC-seq data from seven blood cell types were downloaded from https://github.com/caleblareau/singlecell_bloodtraits/tree/master/data/bulk/ATAC/narrow peaks. The BECs ATAC-seq data are available as supplementary material from the original publication: https://doi.org/10.1038/s42003-020-01411-4. The PCHi-C datasets are available as supplementary material from the original publications: https://doi.org/10.1016/j.cell.2016.09.037 (blood immune cells), https://doi.org/10.1038/s42003-020-01411-4 (BECs), and https://doi.org/10.1038/s42003-020-01411-4 (bulk lung). The ABC models are available on Engreitz Lab’s website: https://www.engreitzlab.org/resources/. GTEx V8 eQTLs were downloaded from https://www.gtexportal.org/home/downloads/adult-gtex/qtl. DICE eQTLs were downloaded from https://dice-database.org/downloads#eqtl_download. OneK1K single-cell eQTLs in peripheral blood mononuclear cells were downloaded from https://onek1k.org/. Single-cell eQTLs in lung were downloaded from https://www.ncbi.nlm.nih.gov/geo/query/acc.cgi?acc=GSE227136. All analysis code is available on GitHub: https://github.com/ez-xyz/asthma_finemapping.

## Abbreviations

GWAS: genome-wide association study
AOA: adult-onset asthma
COA: childhood-onset asthma
CRE: *cis*-regulatory element
MPRA: massively parallel reporter assay
BEC: bronchial epithelial cell
eQTL: expression quantitative trait locus
LD: linkage disequilibrium
TWAS: transcriptome-wide association study
sQTL: splicing quantitative trait locus
caQTL: chromatin accessibility quantitative trait locus
meQTL: DNA methylation quantitative trait locus
UKB: UK Biobank
SNP: single nucleotide polymorphism
S-LDSC: stratified LD score regression
OCR: open chromatin region
snATAC-seq: single-nucleus ATAC-seq
SuSiE: sum of single effects
PIP: posterior inclusion probability
ePIP: element PIP
PCHi-C: promoter capture
Hi-C ABC: activity-by-contact
GO: Gene Ontology
CPM: counts per million
TSS: transcription start site
TLR: toll-like receptors
OX40L: OX40 ligand
RV: rhinovirus

## Data Availability

This study uses genotype and phenotype data from the UK Biobank under application number 44300. Access to UK Biobank resource is available with application at http://www.ukbiobank.ac.uk. Summary statistics of AOA and COA GWAS performed with UK Biobank version 3 genotypes will be made available prior to publication of the manuscript. The sequencing data generated in this study were deposited in EMBL-EBI's Array Express database (https://www.ebi.ac.uk/biostudies/arrayexpress) under accession numbers E-MTAB-14267 (ASMCs ATAC-seq), E-MTAB-14273 (BECs MPRA), and E-MTAB-14295 (ASMCs PCHi-C). The snATAC-seq data from 18 lung cell types were downloaded from https://www.lungepigenome.org. The ATAC-seq data from seven blood cell types were downloaded from https://github.com/caleblareau/singlecell_bloodtraits/tree/master/data/bulk/ATAC/narrowpeaks. The BECs ATAC-seq data are available as supplementary material from the original publication: https://doi.org/10.1038/s42003-020-01411-4. The PCHi-C datasets are available as supplementary material from the original publications: https://doi.org/10.1016/j.cell.2016.09.037 (blood immune cells), https://doi.org/10.1038/s42003-020-01411-4 (BECs), and https://doi.org/10.1038/s42003-020-01411-4 (bulk lung). The ABC models are available on Engreitz Lab website: https://www.engreitzlab.org/resources/. GTEx V8 eQTLs were downloaded from https://www.gtexportal.org/home/downloads/adult-gtex/qtl. DICE eQTLs were downloaded from https://dice-database.org/downloads#eqtl_download. OneK1K single-cell eQTLs in peripheral blood mononuclear cells were downloaded from https://onek1k.org/. Single-cell eQTLs in lung were downloaded from https://www.ncbi.nlm.nih.gov/geo/query/acc.cgi?acc=GSE227136. All analysis code is available on GitHub: https://github.com/ez-xyz/asthma_finemapping.

## Acknowledgements

The authors would like to acknowledge William Wentworth-Sheilds for assistance with data curation, Dr. Kevin Luo for assistance with the mapgen R package, Dr. Milton Pividori for assistance with creating the Miami plot, and Dr. Caleb Lareau for helpful discussions about the ATAC-seq data from blood cells. The authors gratefully acknowledge the Gift of Hope Organ and Tissue Donor Network and their donors and their families for providing tissues used for this study.

## Author information

### Authors and Affiliations

Department of Human Genetics, University of Chicago, Chicago, IL, 60637, USA

Xiaoyuan Zhong, Robert Mitchell, Christine Billstrand, Emma Thompson, Noboru J. Sakabe, Ivy Aneas, Isabella M. Salamone, Jing Gu, Marcelo A. Nóbrega, Xin He & Carole Ober

Division of Pulmonary and Critical Care Medicine, Department of Medicine, University of Virginia, Charlottesville, VA, 22908, USA Anne I. Sperling

Section of Pulmonary and Critical Care Medicine, Department of Medicine, University of Chicago, Chicago, IL, 60637, USA

Nathan Schoettler

## Contributions

X.Z. harmonized the datasets, performed the computational analyses, interpreted the results, and prepared the manuscript; R.M and E.T. performed ATAC-seq and PCHi-C in ASMCs; C.B. performed luciferase assays in BECs; I.A. performed MPRA in BECs; N.J.S. processed the ATAC-seq, PCHi-C, and MPRA data; J.G. assisted with data harmonization and computational analyses; I.M.S., A.I.S., and N.S. assisted with results interpretation; M.A.N. supervised the functional genomics experiments; X.H. and C.O. supervised the computational analyses; M.A.N., X.H., and C.O. designed the study and interpreted the results. All authors contributed to writing the manuscript. All authors read and approved the final manuscript.

## Corresponding authors

Correspondence to Xiaoyuan Zhong, Marcelo A. Nóbrega, Xin He or Carole Ober.

## Ethics declarations

### Ethics approval and consent to participate

Institutional Review Board (IRB) approval was waived because this research was not deemed to constitute human subject research. For the same reason, consent to participate was not applicable.

### Consent for publication

Not applicable.

