## Additional File 1 for "Integration of functional genomics and statistical fine-mapping systematically characterizes adult-onset and childhood-onset asthma genetic associations"

**Supplementary Methods**

GWAS of AOA and COA

We previously performed GWAS of AOA and COA [1] using version 2 of the UK Biobank [2] (UKB) imputed genotypes. Because some single nucleotide polymorphisms (SNPs) in the version 2 imputed data were later found to have wrong genomic positions, the UKB released version 3 of the imputed data with corrected positions. We thus re-ran the AOA and COA GWAS with version 3 imputed genotypes, following the same quality control and analysis pipeline, and using the same definitions for cases and controls, as in our previous study [1]. The sample included 20,702 AOA cases, 9,613 COA cases, and 308,537 shared controls from UKB White British unrelated individuals. For each GWAS, we ran logistic regression for 9,738,009 autosomal SNPs using the logistic_regression_rows() function in Hail 0.2 [3]. We included sex and the first 10 genetic principal components as covariates.

Chromatin accessibility data harmonization

Blood chromatin accessibility data were obtained from Ulirsch et al. [4], who performed ATAC-seq on seven sorted populations of blood cells. Lung chromatin accessibility data were collected from three sources: Wang et al. [5], who performed single-nucleus ATAC-seq (snATAC-seq) on cells from non-diseased lungs and annotated the cells into 18 cell types; Helling et al. [6], who conducted ATAC-seq on cultured primary bronchial epithelial cells (BECs); and an ATAC-seq dataset generated for this study using cultured primary airway smooth muscle cells (ASMCs). The 27 blood and lung cell types were grouped into five lineages, following the lineage definitions in Wang et al. [5]: Lymphoid (lung B cells, lung T cells, lung NK cells, blood B cells, blood $CD4^+$ T cells, blood $CD8^+$ T cells, blood NK cells); Myeloid (lung macrophage, blood myeloid dendritic cells, blood plasmacytoid dendritic cells, blood monocytes); Epithelial (alveolar type 1 cells, alveolar type 2 cells, pulmonary neuroendocrine cells, lung basal cells, lung ciliated cells, lung club cells, BECs); Mesenchymal (lung matrix fibroblasts 1, lung matrix fibroblasts 2, lung myofibroblasts, lung pericytes, ASMCs); Endothelial (lung arterial cells, lung capillary 1 cells, lung capillary 2 cells, lung lymphatic cells).

#### Functional fine-mapping

We used an empirical Bayesian model, TORUS [7], to integrate functional annotations (i.e., OCRs in cell lineages significantly enriched for AOA/COA heritability) with fine-mapping. Briefly, TORUS first estimates an enrichment parameter for each annotation, which reflects how much more likely a SNP with this annotation is causal compared to a randomly selected SNP. Next, TORUS computes a functional prior for each SNP, based on its overlapped annotations. In general, SNPs that overlapped with annotations enriched at GWAS loci were assigned higher prior probabilities in fine-mapping.

To facilitate fine-mapping, we divided the genome into 1,703 approximately independent LD blocks, calculated using LDetect [8] on 1000 Genomes European populations [9], and selected LD blocks that contained at least one genome-wide significant SNP (significant blocks). We excluded LD blocks spanning the HLA region, which we have fine-mapped in a previous study [cite Selene's paper]. We ran the `susie_rss()` function from susieR R package version 0.12.35 on each significant block, which takes GWAS z-scores and correlations of SNPs in the LD block as input and estimates a posterior inclusion probability (PIP) for each SNP. We used in-sample LD and specified the prior probabilities of individual SNPs using the functional priors computed by TORUS in the first step. We allowed up to 5 causal signals for each LD block, set the level of credible set at 95%, and used a purity threshold of 0.4 to filter credible sets. SNPs with minor allele frequencies greater than 0.01 were included in fine-mapping. For multi-allelic SNPs, we only kept the first instance in GWAS summary statistics.

For comparison, we also performed fine-mapping without using functional information. For this, we ran the `susie_rss()` function with uniform priors while keeping other parameters the same.

<u>Gene score calculation</u>

The gene scores we derived summarize the total genetic evidence supporting the role of a gene as an AOA or COA risk gene. The score of a gene $g$, $S_g$, was the sum of the contributions of all variants linked to $g$. We denote the contribution of the variant $j$ to the gene $g$ as $S_{gj}$. To compute these contributions, we considered exonic variants and non-coding variants in candidate CREs with PIP > 0.1. If $j$ is an exonic variant of $g$, then $S_{gj}$ equals $\mathrm{PIP}_j$, the PIP of the variant $j$ from fine-mapping. If $j$ is a non-coding variant in candidate CRE, we defined a "link score" between the variant $j$ and the gene $g$ as $L_{gj}$ (see below), which measures the strength of evidence supporting this variant-gene pair.

The contribution of variant $j$ to gene $g$ is $S_{gj} = L_{gj} \times \mathrm{PIP}_j$, the link score weighted by the PIP of the variant.

The total link score of a non-coding variant to a gene was the sum of link scores over the four categories of evidence linking a CRE to a gene, described in the Methods section "Linking candidate CREs to target genes". If a category implicated a single target gene for a candidate CRE, then the link score of that gene from that category is 1. When an evidence category suggested multiple target genes for a candidate CRE, the link score of each gene in that category is 1 divided by the number of putative target genes. For a variant in genic region, we additionally nominated its residing gene as a target gene under the distance category if it is not the nearest gene. We defined genes with gene score ≥ 0.95 as high-confidence candidate causal genes for AOA or COA. We restricted this analysis to protein-coding genes.

Identifying the cellular contexts of fine-mapped variants

The goal of this analysis was to identify the cell lineage(s) through which the fine-mapped variants act. If we had full confidence that a SNP was a causal variant, this task would be simply achieved by checking whether the variant overlaps with an OCR in any of the cell lineages. However, given the uncertainty of causal variants, we analyzed all variants within a credible set, while weighting the variants by their PIPs. Specifically, if a SNP overlapped with an OCR in only one lineage, we assigned the PIP of that SNP to that lineage. If the SNP overlapped with an OCR shared by ≥ 2 lineages, then the SNP PIP assigned to lineage $j$, $PIP_j$, was computed as:

$$PIP_j = SNP\ PIP \times \frac{w_j}{\sum_{j=1}^{J} w_j},$$

where $w_j$ is the proportion of SNP heritability explained by OCRs of lineage $j$, estimated using S-LDSC. For each credible set, we divided the total PIP assigned to a lineage by the total PIP of the credible set to determine the proportion of PIP attributed to that lineage. This proportion of a lineage can be interpreted as the probability that this lineage is the cellular context of the causal variant(s) within each credible set.

<u>Testing the enrichment of MPRA enhancers in high-ePIP candidate CREs</u>

We partitioned the candidate CREs harboring SNPs assayed by MPRA in BEC into two sets: an $MPRA^+$ set, which were those that contained SNPs in MPRA-validated enhancer sequences, and an $MPRA^-$ set. We then performed Wilcoxon rank-sum test to compare the ePIP distributions of the two sets of candidate CREs, using the `wilcox.test()` function in R.

<u>Gene ontology enrichment</u>

WebGestalt [10] was used to assess the enrichment of Gene Ontology [11,12] (GO) Biological Process terms among the high-confidence candidate causal genes (gene score ≥ 0.95). Over-representation analysis was performed with “Homo sapiens” as the Organism of Interest, “GO Biological Process noRedundant” as the Functional Database, and all protein-coding genes as reference set. All advanced parameters were kept as default. A set of top enriched GO terms were generated using the weighted set cover algorithm.

ATAC-seq in ASMCs

ATAC-seq data were generated using primary ASMCs from four donors whose lungs were unable to be transplanted at the time of death. Donors were male, self-reported white, non-smokers ranging in age from 45 to 62 years of age. ASMCs at passage 1 or 2 were cultured following the protocol described in Thompson et al. [13]. On the final day of culture, cells were trypsinized and counted manually, then separated into aliquots of 50,000 cells for processing.

ATAC-seq was performed as described by Grandi et al. [14]. Briefly, 50,000 cells were collected at 500 g for 5 min at 4°C and gently resuspended in 50 µl ice-cold lysis buffer (10 mM Tris–HCl pH 7.5, 10 mM NaCl, 3 mM MgCl2, 0.1% NP40, 0.1% Tween-20 and 0.01% digitonin). Lysis proceeded for 3 minutes on ice and was neutralized with 1 ml of ice cold lysis buffer excluding detergents digitonin and NP40. Following neutralization, permeabilized nuclei were pelleted at 500 g for 10 minutes at 4°C and resuspended in 50 µl transposition master mix (25 µl (2X) Illumina TD buffer, 16.5 µl PBS, 5 µl H2O, 0.5 µl 1% digitonin, 0.5 µl 10% Tween-20, and 2.5 µl Illumina TDE1 enzyme (Tn5 transposase)) and incubated for 30 minutes at 37°C on a thermomixer set to 1000 rpm. Tagmentation was terminated and DNA collected using Zymo Clean and Concentrator-5 following the manufacturers recommendations. Libraries were barcoded with 5 cycles of preamplification, and the PCR reaction was paused for quantification using NEBNext

Library Quant Kit for Illumina. Additional cycles were ran as needed to produce at least 10 nM of final library. Purified ATAC-seq libraries were again quantified, diluted to 8 nM and assayed for QC on an Agilent 2100 Bioanalyzer High Sensitivity Chip. Diluted final libraries were mixed and sequenced on an Illumina Nova-Seq using paired end 50 bp reads with a target depth of ~50M reads/sample.

ATAC-seq reads were aligned with bowtie2 [15] version 2.3.4.3 with parameters `-X 2000 --fr --no-discordant --very-sensitive-local`. Reads with mapping quality lower than 10 were discarded. Peaks were called using MACS2 [16] version 2.2.7.1 with parameters `--llocal 20000 --shift -100 --extsize 200 -q 0.05`. Peaks overlapping hg19 coordinates blacklisted by ENCODE [17] were excluded.

PCHi-C in ASMCs

Primary ASMCs obtained from the source as described above were isolated, cryopreserved, and cultured as described previously [13]. We performed PCHi-C using three replicates each from two non-asthmatic donors, as described previously [18,19]. HiCUP [20] version 0.5.9 was used to map PCHi-C reads to the genome and remove technical artifacts. CHiCAGO [21] version 1.20.0 was run on filtered reads to detect significant interactions, defined as CHiCAGO score > 5.

MPRA in BECs

We selected the lead SNPs at individual loci from our previously published AOA and COA GWAS [1]. LDproxy [22] was then used to identify a list of 2,034 SNPs within 500 kb of and LD $r^2 > 0.8$ with the lead SNP. MPRA was performed using these SNPs in 16HBE14o-, a human BEC line, as described previously [23,24]. MPRA data were analyzed following procedures described in Ulirsch et al. [25]. First, an enhancer activity was calculated for each construct by obtaining the log of the counts per million (CPM) in each sample and doing: RNA log(CPM) - DNA log(CPM) to correct RNA counts of each construct for differences in library representation. RNA log(CPM) lower than -2 and DNA log(CPM) lower than 0 were removed based on the kernel densities of the counts that showed high variance of low counts. Activities were then quantile normalized and centered around the median (normalized activity - median of all activities). To identify constructs that acted like enhancers, i.e. whose activities were higher than background, we compared the activity of each construct with all the other activities using the `wilcox.test()` function in R and corrected the one-tailed p-values using the FDR method in the `p.adjust()` R function. Constructs whose tests resulted in an adjusted p-value lower than 0.05 were considered putative enhancers in each replicate. Only constructs that were considered enhancers in at least 3 out of 4 replicates were accepted in the final list of enhancers. Differences of activity between two constructs bearing different alleles were tested for those constructs for which at least one of the alleles was considered an enhancer using a two-tailed Wilcoxon test comparing the activities of all barcodes for each construct.

<u>Luciferase assays in BECs</u>

Luciferase assays were performed in a human BEC line 16HBE14o- (Millipore Cat. No. SCC150). The cells were grown on Fibronectin/Collagen/BSA ECM coated flasks and 24 well plates. The growth media consisted of Alpha MEM (Sigma Cat. No. M2279), 10% FBS (Sigma Cat. No. ES-009-B), 2 mM L-Glutamine (Sigma Cat. No. TMS-002-C), and 1X Penicillin-Streptomycin Solution (Sigma Cat. No. TMS-AB2-C). Cells were incubated at 37°C in a humidified incubator with 5% CO2 and passaged at 90-95% confluency with Trypsin-EDTA solution (Sigma Cat. No. T3924).

100,000 16HBE14o- cells/well were seeded with growing media on a 24 well plate. Transfections were done at 80% cell confluency using Lipofectamine LTX reagent with PLUS™ reagent (Life Technologies Cat. No. 15338100) in Opti MEM I (Life Technologies Cat. 31985062) media and incubated at 37°C for 24 hours.

Each plasmid was co-transfected with a *Renilla* plasmid as an internal control. The individual plasmids were transfected in triplicate wells for each experimental run. Plasmids were constructed using pGL4.23 (Promega Cat. No. E8411) for the enhancer assay or pGL4.10 (Promega Cat. No. E6651) for the promoter assay as the backbone for the various regions tested. For each candidate CRE, DNA sequence +/- 500 bp to the SNP with the highest PIP were obtained from UCSC Genome Browser [26] hg19 assembly of the human genome. All genome-wide significant SNPs within the +/- 500 bp window were extracted from AOA/COA GWAS summary statistics, and the observed haplotypes among 1000 Genomes European populations were obtained using LDhap [22]. The constructs were manufactured by GenScript. A desert DNA plasmid was used as a negative control and pGL3 SV40 as a positive control. A Dual-Luciferase reporter assay (Promega Cat. No. E1910) was used to measure luciferase activity. 20 µl of lysed cells from each well were placed in a 96 well optical plate. A Promega Luminometer plate reader using the DualGlo program first adding 100 µl LAR II to measure firefly luciferase activity then adding 100 µl Stop & Glo to measure the *Renilla* luciferase activity.

For each reading well, we calculated the ratio between firefly luciferase activity and *Renilla* luciferase activity. Hereafter, this ratio is referred to as the normalized luciferase activity. The normalized luciferase activities were averaged across the triplicate wells within each technical replicate. Three technical replicates were conducted for each candidate at first, and more technical replicates were performed to assess haplotype-specific effects if enhancer or promoter activities were suggested by the first three technical replicates. The $\log_2$ fold changes of average normalized luciferase activity between different haplotypes and between haplotypes and control were calculated for each technical replicate. To determine the statistical significance of haplotype-specific effect on enhancer/promoter activity, a two-tailed, paired t-test was performed to compare the $\log_2$ fold changes using `t.test()` function in R.

### Supplementary Figures

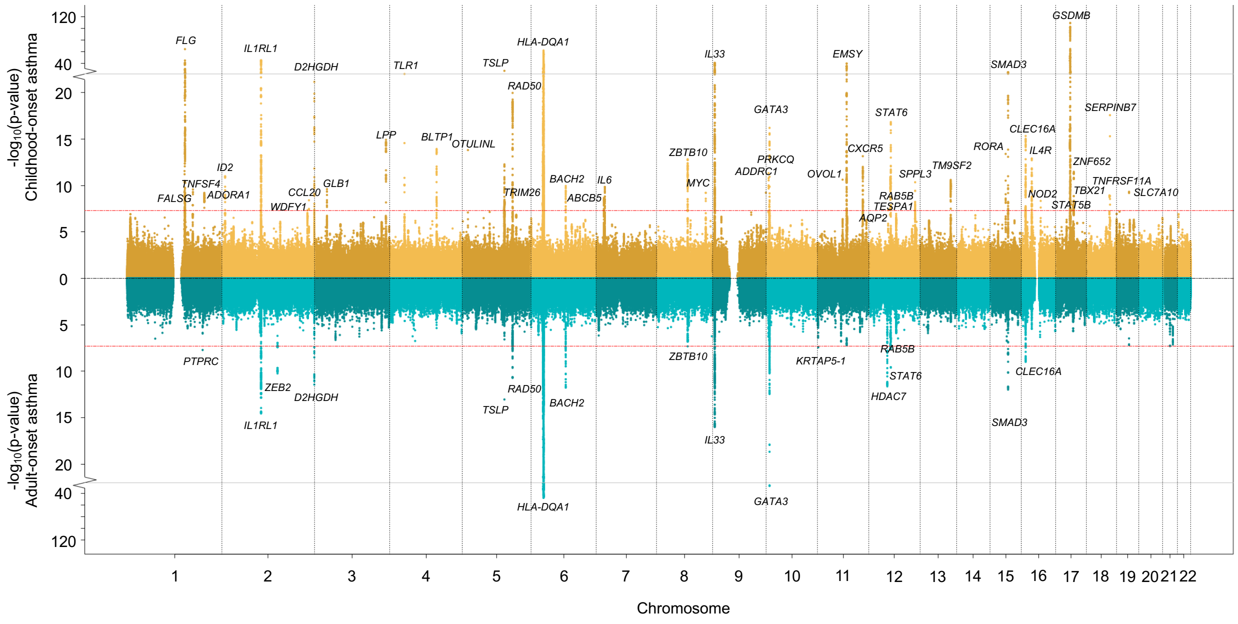


**Fig. S1.** Miami plot for adult-onset asthma (bottom) and childhood-onset asthma (top) GWAS. Red dashed line marks the genome-wide significance threshold (p-value < $5 \times 10^{-8}$). Genome-wide significant loci were labelled by the nearest gene to the most significant SNP.

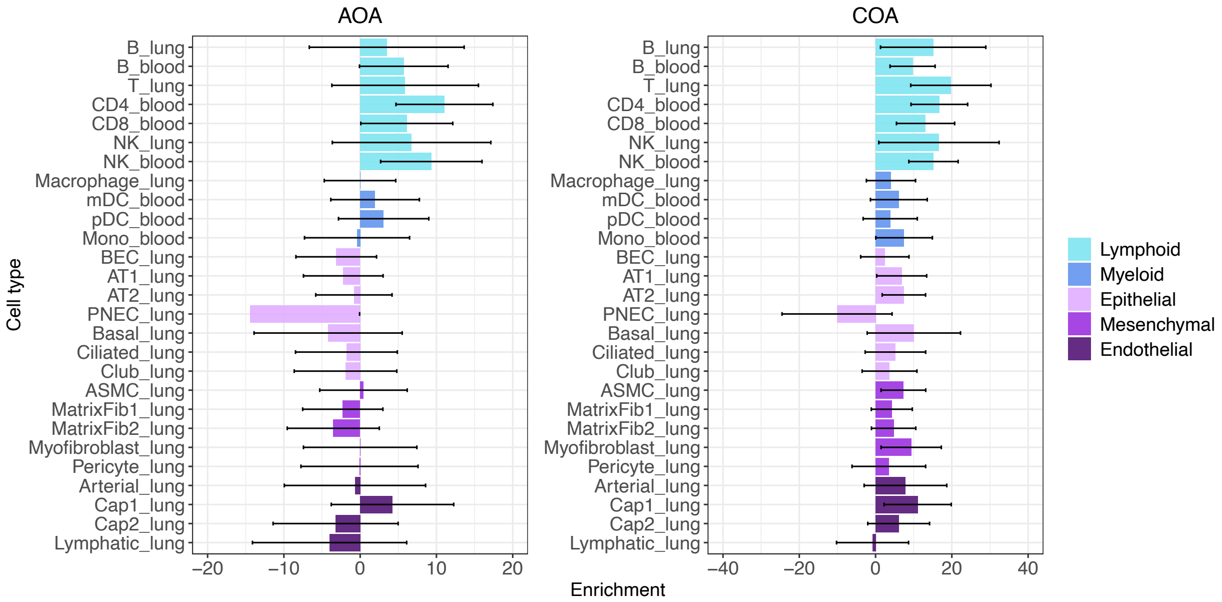


**Fig. S2.** S-LDSC heritability enrichment estimates for 7 blood and 20 lung cell types for adult-onset asthma (left panel) and childhood-onset asthma (right panel). The horizontal bars are confidence intervals (+/- 2 standard errors).

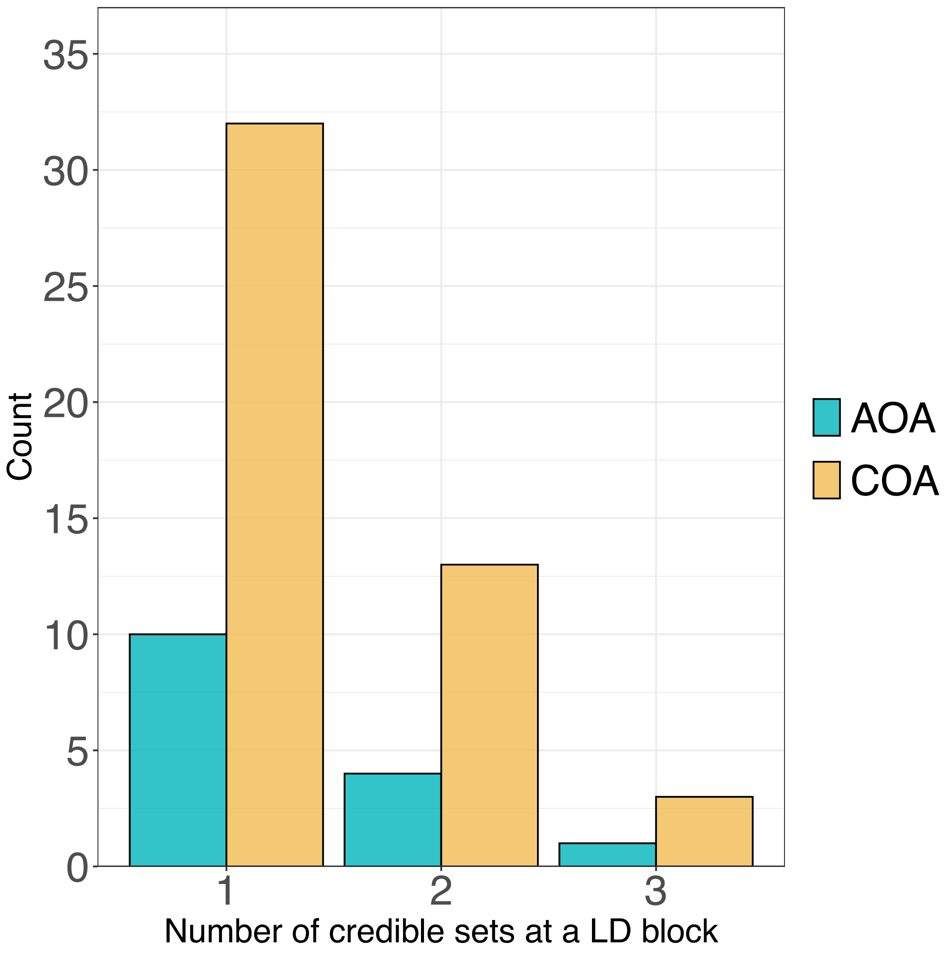


**Fig. S3.** Distribution of the number of credible sets at an LD block for adult-onset asthma and childhood-onset asthma.

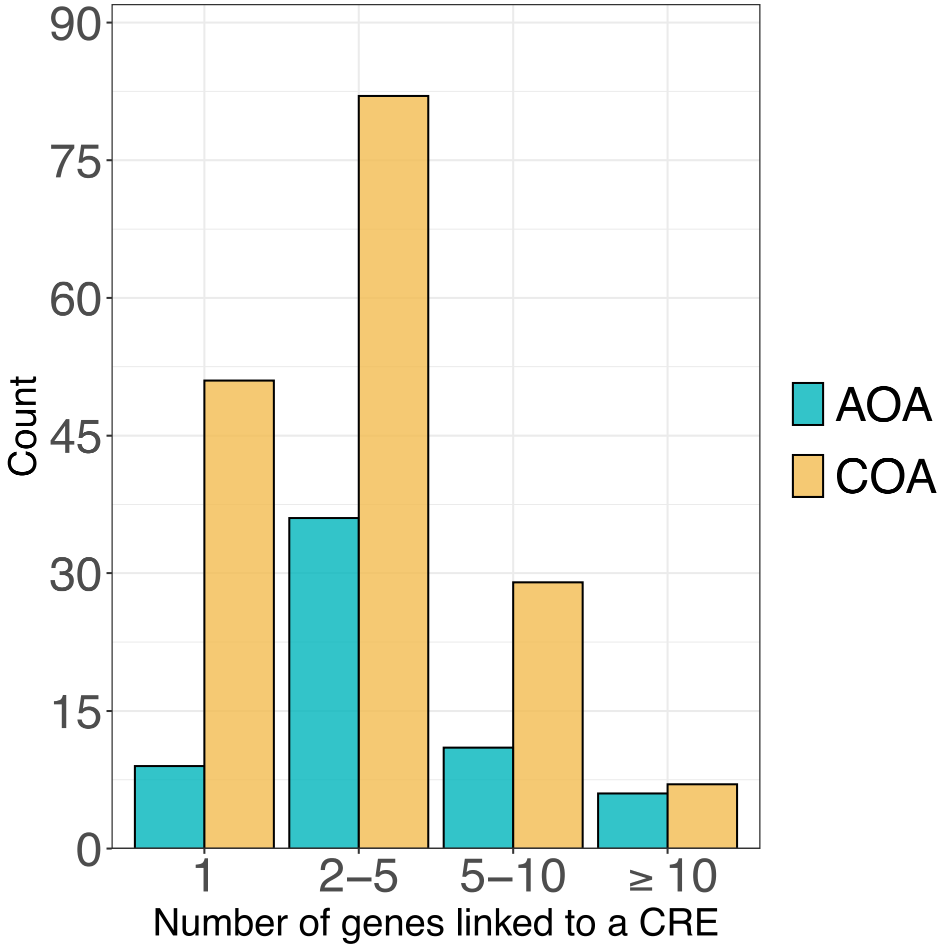


**Fig. S4.** Distribution of the number of genes linked to a candidate *cis*-regulatory element (CRE) with nonzero ePIP. Genes were assigned to CREs based on distance (i.e., the nearest gene), PCHi-C, ABC, and eQTL data.

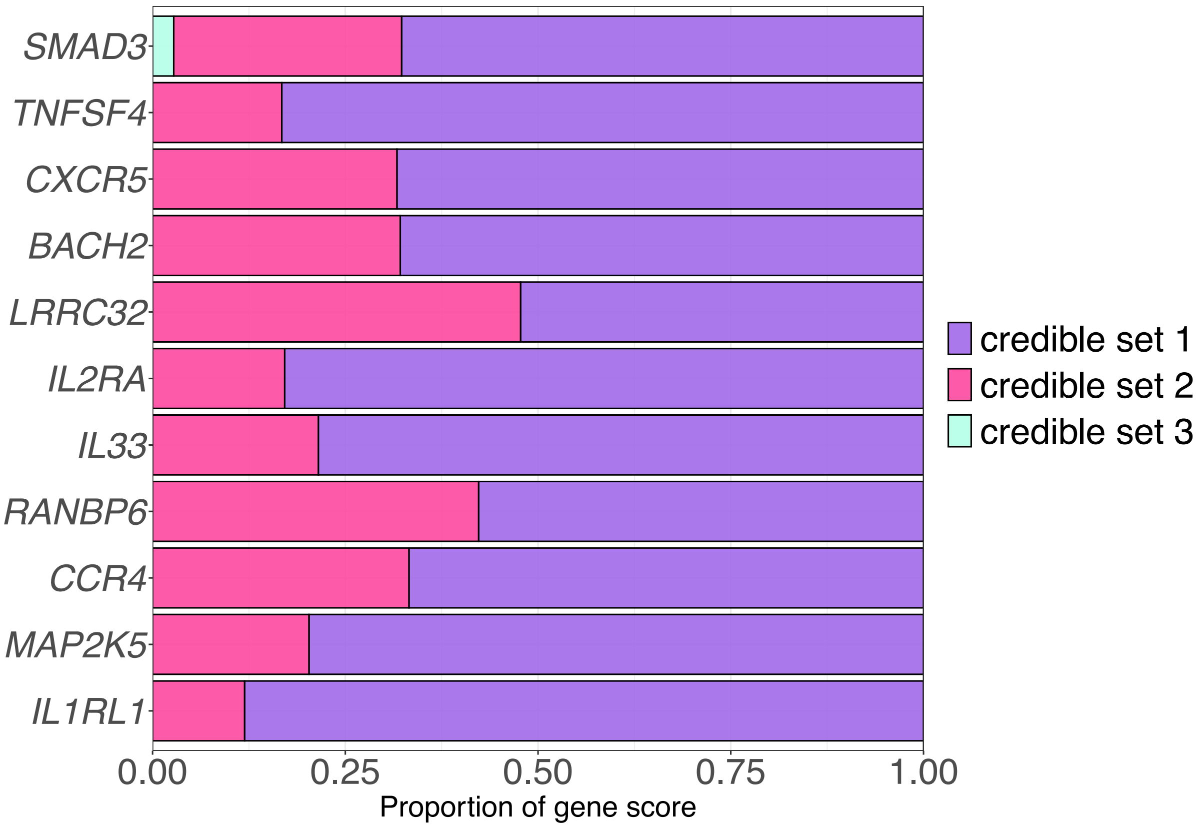


**Fig. S5.** Proportions of gene scores attributed to individual credible sets for COA high-confidence candidate causal genes (gene score ≥ 0.95) that were targeted by more than credible sets. Genes were plotted only if the two most significant credible sets each accounting for at least 10% of the gene score.

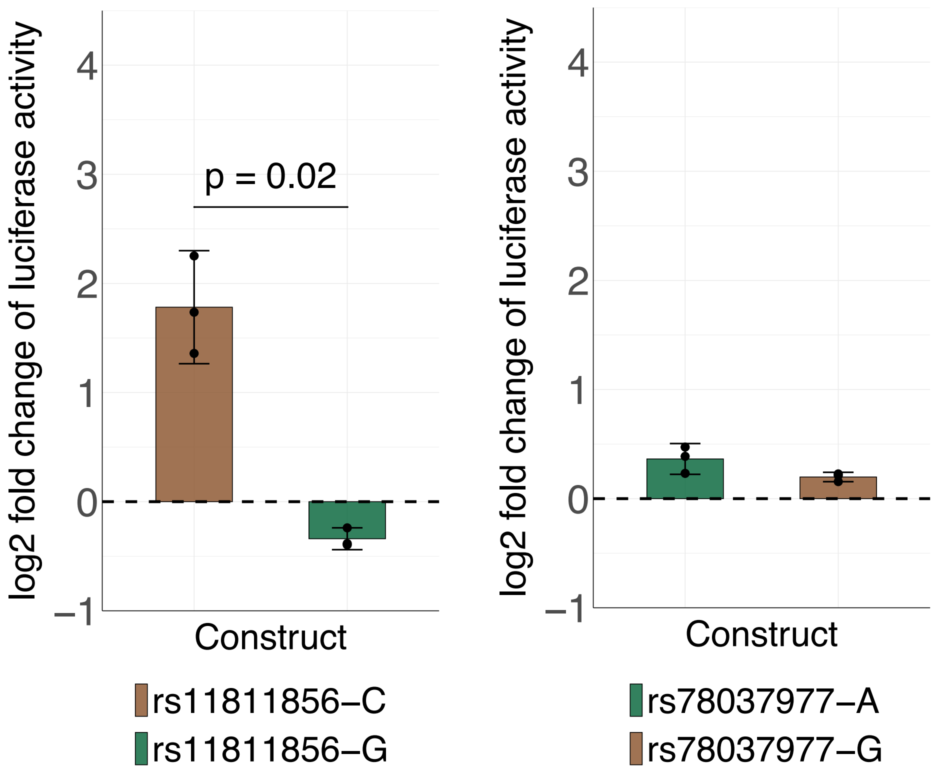


**Fig. S6.** Luciferase assay results in bronchial epithelial cells for the sequences containing rs11811856 (left) and rs78037977 (right). Different colors correspond to different constructs, and the constructs containing asthma risk allele of the SNP with the highest-PIP are colored green. For each construct, the log2 fold changes of the average normalized luciferase activity relative to the control (dashed line at 0) are plotted across experimental replicates. The height of the bar shows the mean log2 fold change, and the confidence intervals are plotted as +/- 2 standard errors from the mean. The p-values were computed using two-tailed, paired t-test. N = 3 experiments for each candidate SNP.

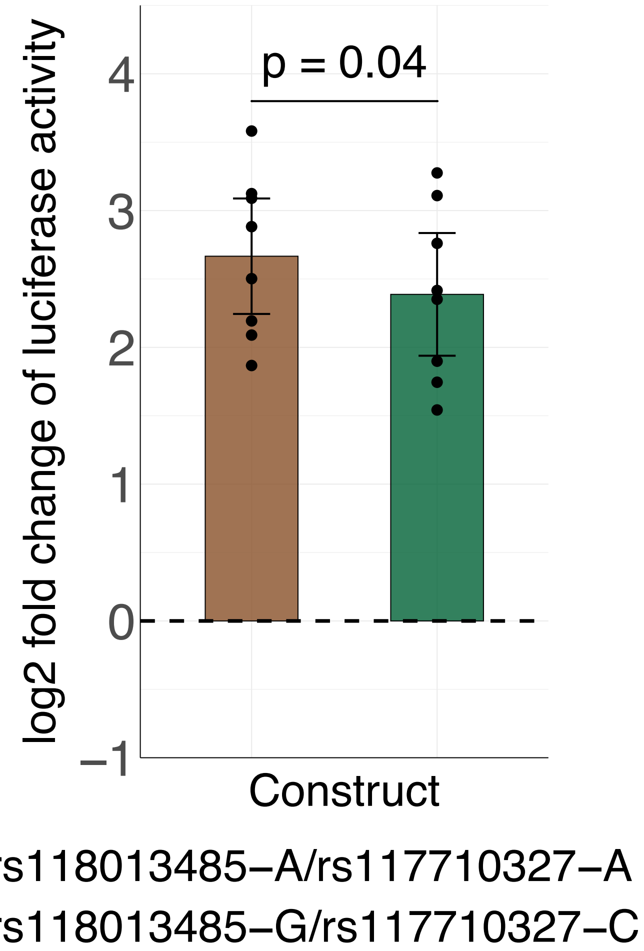


**Fig. S7.** Luciferase assay results in bronchial epithelial cells for the enhancer sequence containing rs118013485/rs117710327 (N = 8 experiments). See Fig. S6 figure legend.

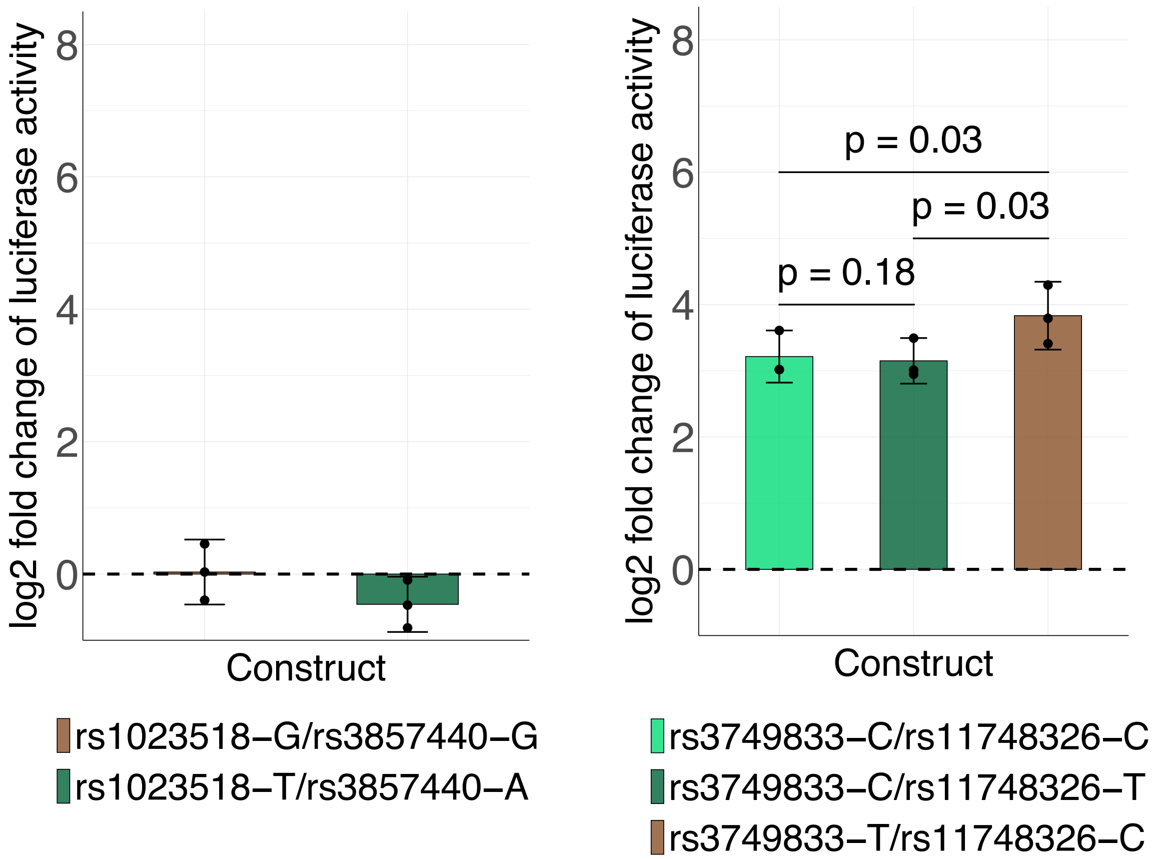


**Fig. S8.** Luciferase assay results in bronchial epithelial cells for the sequences containing rs1023518/rs3857440 (left) and rs3749833/rs11748326 (right). N = 3 experiments for each candidate SNP. See Fig. S6 figure legend.

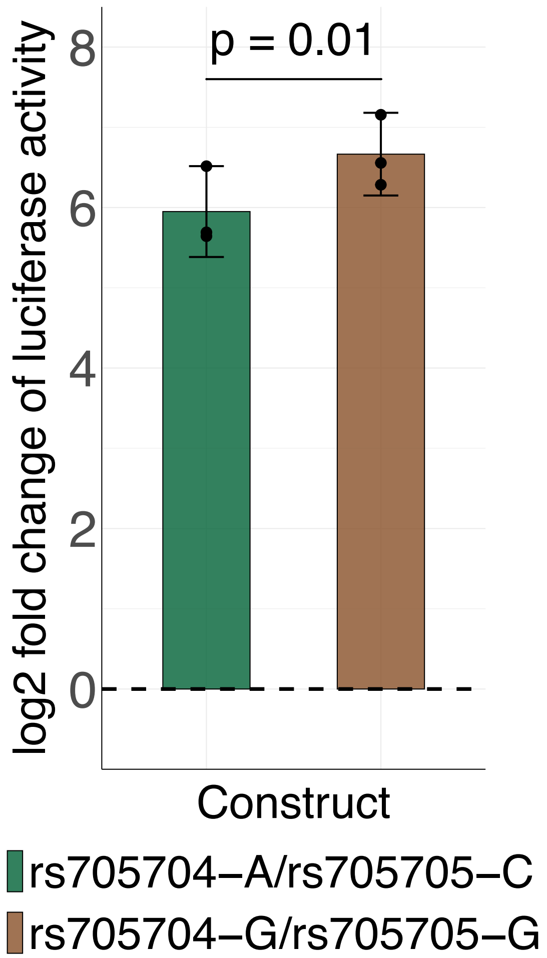


**Fig. S9.** Luciferase assay results in bronchial epithelial cells for the sequences containing rs705704 and rs705705 (N = 3 experiments). See Fig. S6 figure legend.
